# Enhancing social inclusion through the implementation of evidence-based digital health interventions for mental health and alcohol/other drug use problems in the wake of COVID-19

**DOI:** 10.1101/2023.09.20.23295867

**Authors:** Frances Kay-Lambkin, Jane Rich, Dara Sampson, Louise Thornton, Milena Heinsch, Kate Filia, Brian Kelly, Alan Weiss, Murray Wright, Danielle Simmonette, Maree Teesson

## Abstract

At any one time, over 900 million people globally experience a mental disorder (including alcohol/other drug use disorders, Whiteford et al., 2013), and this is increasing by about 3% each year (ABS, 2018). Adding to these challenges, the COVID-19 pandemic presents clear risks for a substantial decline in global mental health. Preliminary evidence points towards an overall rise in symptoms of anxiety and coping responses to stress (Holmes et al., 2020), including increased drug and alcohol use amongst the general population. The greatest mental health impacts of the COVID-19 pandemic will be felt, however, by those who are already most marginalised and people with pre-existing mental health and substance use disorders, who have a higher susceptibility to stress than the general population (Yao et al., 2020). eCliPSE is an online clinical portal developed by CI Professor Frances Kay-Lambkin in partnership with the research team and the NSW Ministry of Health to facilitate access to evidence-based ehealth treatments for mental health and alcohol/other drug [AOD] use problems. However, since the testing of eCliPSE in 2017, uptake of this tool via clinician referral has been low, and no clear models existed for digital treatment integration into health services (Batterham et al., 2015). There are very few examples in the available literature of successful implementation of digital interventions in clinical services, and many failures (Mohr et al., 2017). In response to this, our team has developed an evidence-informed Integrated Translation and Engagement Model (ITEM) to drive the uptake of digital therapeutics into mental health and alcohol/other drug services across NSW. Based on the latest evidence for effective implementation, a consideration of individual, social, environmental, and structural factors, the ITEM synthesises diverse theoretical approaches into a coherent, integrated model. The pandemic has highlighted (and exacerbated) social inequities in relation to the prevalence of mental illness, as well as treatment options. Technology has the potential to respond to this challenge, but Australia lags behind the rest of the world in implementing sustainable, effective digital tools into health service delivery. Additionally, no tool currently exists for the evaluation of dual diagnosis capability of digital programs.

## 2 INTRODUCTION

### 2.1 Background and rationale

At any one time, over 900 million people globally experience a mental disorder (including alcohol/other drug use disorders, Whiteford et al., 2013), and this is increasing by about 3% each year (ABS, 2018). Adding to these challenges, the COVID-19 pandemic presents clear risks for a substantial decline in global mental health. Preliminary evidence points towards an overall rise in symptoms of anxiety and coping responses to stress (Holmes et al., 2020), including increased drug and alcohol use amongst the general population. The greatest mental health impacts of the COVID-19 pandemic will be felt, however, by those who are already most marginalised and people with pre-existing mental health and substance use disorders, who have a higher susceptibility to stress than the general population (Yao et al., 2020).

eCliPSE is an online clinical portal developed by CI Professor Frances Kay-Lambkin in partnership with the research team and the NSW Ministry of Health to facilitate access to evidence-based ehealth treatments for mental health and alcohol/other drug [AOD] use problems. However, since the testing of eCliPSE in 2017, uptake of this tool via clinician referral has been low, and no clear models existed for digital treatment integration into health services (Batterham et al., 2015). There are very few examples in the available literature of successful implementation of digital interventions in clinical services, and many failures (Mohr et al., 2017). In response to this, our team has developed an evidence-informed Integrated Translation and Engagement Model (ITEM) to drive the uptake of digital therapeutics into mental health and alcohol/other drug services across NSW. Based on the latest evidence for effective implementation, a consideration of individual, social, environmental, and structural factors, the ITEM synthesises diverse theoretical approaches into a coherent, integrated model.

The pandemic has highlighted (and exacerbated) social inequities in relation to the prevalence of mental illness, as well as treatment options. Technology has the potential to respond to this challenge, but Australia lags behind the rest of the world in implementing sustainable, effective digital tools into health service delivery. Additionally, no tool currently exists for the evaluation of dual diagnosis capability of digital programs.

### 2.2 Research Aims

This study will:

I. Explore the experiences, attitudes, and perspectives of digital health tools among Ramsay mental health and AOD staff and service users,
II. Investigate mental health, AOD and social equity factors in a sample of service users from Ramsay and non-Ramsay services (i.e., public, and private) in the Hunter New England and North Sydney catchment areas,
III. Redevelop the ITEM to include a social inclusion component for the implementation of digital tools into services that account for and address factors of social exclusion, and
IV. Redevelop the Dual Diagnosis Capability Assessment Tool (DDCAT) to include an eHealth component (eHCAT) for the assessment of dual diagnosis capability of digital interventions.

### 2.3 Study Design

To address the aims of the study, a concurrent mixed-method design will be employed. The quantitative component of the study will include a targeted survey assessing mental health and AOD symptomology, social exclusions, digital health use, treatment access and history, and needs, values, and preferences relating to digital health use. The qualitative component of the study will include ono-on-one interviews with clinicians and patients of Ramsay and non-Ramsay mental health and AOD services to scope attitudes of, preferences for, and experience with, digital mental health.

## 3 SURVEY METHODS

### 3.1 Study Setting

Patients of Ramsay (Ramsay Psychology, St Leonards; and Ramsay Lakeside Clinic, Warners Bay) and non-Ramsay services in the Hunter New England (HNE) and North Sydney areas (services to be confirmed).

### 3.2 Recruitment

Patients of Ramsay and non-Ramsay services will be invited by email and text message. Contact with potential survey respondents of the Ramsay services will be facilitated chief investigators Professor Murray Wright (Psychiatrist at Ramsay Psychology, St Leonards) and Associate Professor Alan Weiss (Director of Ramsay Lakeside Clinic, Warners Bay). Networks of the research team within NSW Health will facilitate access to the Non-Ramsay Hunter New England sites.

### 3.2 Eligibility Criteria

Participants will be considered to be eligible for this study if they are:

- Aged 18 years and older
- Service user of Ramsay Psychology, St Leonards, or and Ramsay Lakeside Clinic, Warners Bay
- Can read, understand, and write English Language
- Can give informed consent to participate

Participants will be considered ineligible for this study if they do not meet the conditions of the inclusion criteria.

### 3.3 Sample Size

A minimum of 100 respondents will be recruited for the online survey to ensure representation of social exclusion and mental health symptomology.

### 3.5 Consent

Informed e-Consent will be elicited from respondents in the online survey, hosted by REDCap data collection software. Upon clicking the link in the social media campaign, potential respondents will first be provided with the information statement and eligibility questions, before being able to provide their consent to participate.

### 3.6 Data Collection

#### 3.6.1 Methodology

Targeted social media campaign will elicit responses from residents of HNE and North Sydney catchment areas. The campaign with include a brief description of the study and provide a link to the online survey. When redirected to the online survey from the social media campaign, potential respondents will first be provided with the information statement, eligibility questions, and e-Consent form. Once informed consent to participate has been provided, respondents will be directed to the online survey, which will take approximately 20 minutes to complete.

#### 3.6.2 Incentives

At completion of the online survey, participants will have the opportunity to go into a draw to win one of five Coles shopping e-gift voucher. If the respondent consents to go into the draw, they will provide an email address for the distribution of the voucher upon winning the draw. At the completion of the survey data collection, five of the respondents consenting to go into the draw will be randomly chosen to receive an e-gift voucher. The five winning respondents will be notified by email with the accompanying voucher.

### 3.7 Ethical Considerations

It is acknowledged by the research team that it is possible for the respondent to become distressed or upset while answering questions relating to mental health and substance use during completion of the survey. If discomfort is experienced, participants may withdraw their consent to and cease participation at any time. Additionally, a list of support services will be provided both at the completion of the survey and in the instance of survey termination and are also provided in the information statement.

### 3.8 Data Analysis

Descriptive statistics will characterise demographics of the sample. Internal reliability of UTAUT-2 will be determined using Cronbach’s alpha coefficient (α). Analysis of variance (ANOVA) will explore the social, and clinical factors related to social inclusion, DASS21, AUDIT, CUDIT, and SDS scores and self-reported treatment access and history as a function of demographics. Network analysis will map the interplay between these factors, providing insights into the structure of relationships between them.

## 4 INTERVIEW METHODS

### 4.1 Study Setting

i. Ramsay Lakeside Clinic, Warners Bay Private Hospital, Warners Bay
ii. Ramsay Psychology, Ramsay Clinic Northside, St Leonards
iii. Hunter New England NSW Health Service (TBC)
iv. North Sydney NSW Health Service (TBC)

### 4.2 Recruitment

Members of the research team will contact service clinicians of Ramsay and non-Ramsay sites by email to invite them to participate in the telephone interview.

Invitations to service users will be facilitated via service managers and clinicians of the Ramsay and non-Ramsay sites via blind carbon copy function (Bcc) email for anonymity or text message.

Both the service user and clinician email invitations will include the interview information statements,as well as a link to a brief online survey that includes the e-Consent and the online interview booking system. If no available interview times are suited to the service users or clinicians, they can indicate this in the survey and a member of the research team will contact them directly to organise a suitable time.

### 4.3 Eligibility Criteria

Participants will be considered eligible for this study if they are:

- Any clinicians or service users from the mental health and AOD Ramsay and non-Ramsay research sites (confirmed after ethics and SSA approval)
- At least 18 years of age
- Can read, understand, and write English Language
- Can give informed consent to participate and have capacity to participate in the interview

Participants will be considered ineligible for this study if they do not meet the conditions of the inclusion criteria.

### 4.5 Sample Size

Interviews: 20 clinicians and 20 service users across Ramsay and non-Ramsay mental health and AOD services (40 total) will be recruited representing the following:

|  | Ramsay | Non-Ramsay | Total |
| --- | --- | --- | --- |
| Mental Health | Service user = 5 | Service user = 5 | 10 |
|  | Clinician = 5 | Clinician = 5 | 10 |
| AOD | Service user = 5 | Service user = 5 | 10 |
|  | Clinician = 5 | Clinician = 5 | 10 |
|  |  |  | 40 |

### 4.6 Consent

Informed e-Consent for both service users and clinicians will be elicited in the brief online survey, hosted by REDCap data collection software. Upon clicking the link in the email invitation or text, potential respondents will first be provided with the information statement and eligibility questions, before being able to provide their consent to participate.

### 4.7 Data Collection

#### 4.7.1 Methodology

We will recruit 20 clinicians and 20 service users across Ramsay and non-Ramsay sites (40 total). Interview questions will ask clinicians about their experiences, attitudes, and perspectives in relation to digital health implementation within their organisation; and interviews will ask service users about their similar experiences and perspectives of new digital health innovations, their use and referral to these. Data from these interviews will be used to develop the eHCAT component of the DDCAT for assessment of eHealth capability.

#### 4.7.2 Incentives

Service users who participate in the telephone interview will be reimbursed with a $20 Coles e-Gift voucher via email, as recognition of their time and contribution. Clinicians will not be offered reimbursement.

### 4.8 Ethical Considerations

The risks associated with participating in the interviews are very low. No personal or sensitive questions will be asked of clinicians or service users, and the research will not impact their relationship with each other, or the organisation (including access to its services) in any way. Furthermore, participants may withdraw their consent at any time, and this will also not affect their employment/relationship with, or access to, an organisation, not with the University of Newcastle.

The researchers undertaking both the observation and interviews are social workers and psychologists with clinical and research experience with people who experience mental health and/or substance use concerns. In the instance that a service user participant becomes upset during the interview, the researcher will ask them if they wish to continue and remind them that they can ask to stop at any time. If a service user participant chooses to withdraw from the interview, they will be provided with additional support information where appropriate. These included descriptions and contact information for Lifeline (24-hour help line with mental health professionals) and Beyond Blue (24-hour help line with mental health professionals and 3pm-midnight online chat function). In the rare case that a service user participant becomes significantly distressed during the interview, the researcher may obtain explicit verbal consent to put them in touch with their support clinician at the research site for additional support.

### 4.9 Data Analysis

Interviews: thematic analysis of the interview transcripts will be conducted using Brawn and Clarke’s six-step approach (Braun & Clarke, 2006; Braun & Clarke, 2022). A combination of manual and electronic coding and analysis methods will be used, depending on independent researcher preference. Transcripts will initially be read multiple times to facilitate immersion in the data and conduct initial coding, prior to uploading transcripts into NVivo for electronic coding. All data analysis and interpretation will be undertaken by members of the research team who have extensive experience in qualitative research processes.

## 5 DATA STORAGE AND MANAGEMENT

All information and data collected through the research study will be stored in accordance with the University of Newcastle’s safe data storage requirements, using the password protected data storage solution OneDrive. Any potential identifiers will be removed and replaced with pseudonyms and codes. Data will only be accessible by the research team and will be stored for a minimum of five years on the University of Newcastle’s OneDrive secure server. Any hard copies of data, including participant information statements and consent forms, will be scanned, and saved before being securely destroyed. Observation notes and audio recordings of interviews will also be stored on the University of Newcastle’s OneDrive secure server. At the appropriate time, all data – including the secure storage of any names and contact details (phone numbers, email addresses, etc.) – will be securely destroyed in line with the University of Newcastle’s research policy provisions.

## 6 REPORTING AND DISSEMINATION

Findings may be presented in academic publications, journals, and conference proceedings; however, individual participants will not be named or identified in any reports or resources arising from the project. Non-identifiable data may be shared with other parties to encourage scientific scrutiny and to contribute to further research and public knowledge, or as required by law. Findings will also be used to develop recommendations in a final report to the funding body.

## Data Availability

This submission contains a protocol but data collection has not commenced.

